# A Phase III Randomized Clinical Trial: The Impact of Paclitaxel-Based Neoadjuvant Laparoscopic Hyperthermic Intraperitoneal Chemotherapy (NLHIPEC) Followed by Sequential Intravenous Chemotherapy in Advanced High-Grade Serous Ovarian Cancer Patients - Interim Analysis of Safety and Immediate Efficacy from the C-HOC Trial

**DOI:** 10.1101/2023.09.22.23295986

**Authors:** Qun Wang, Hua Liu, Yuhong Shen, Lifei Shen, Weiwei Feng

## Abstract

**Objective:** This study evaluates the potential superiority of combining paclitaxel-based neoadjuvant laparoscopic hyperthermic intraperitoneal chemotherapy (NLHIPEC) with sequential intravenous neoadjuvant chemotherapy over intravenous neoadjuvant chemotherapy (IV NACT) alone in Chinese patients with Federation of Gynecology and Obstetrics (FIGO) stage IIIC-IVB high-grade serous ovarian/fallopian tube carcinoma (HGSOC). This interim analysis focuses on the safety and immediate efficacy of both regimens to determine the feasibility of a planned phase III trial.

**Methods:** In a single-center, open-label, phase III trial, FIGO stage IIIC-IVB HGSOC patients (FAGOTTI score ≥8 during laparoscopic exploration) unsuitable for optimal cytoreduction in primary debulking surgery (PDS) were randomized 2:1 during laparoscopic exploration. The NLHIPEC group received one cycle of intraperitoneal neoadjuvant laparoscopic hyperthermic intraperitoneal chemotherapy (paclitaxel) followed by three cycles of intravenous chemotherapy (paclitaxel plus carboplatin), while the IV NACT group received only three cycles of intravenous chemotherapy. Both groups subsequently underwent interval debulking surgery (IDS). This partial analysis focuses on comparing adverse effects of chemotherapy, postoperative complications, and pathological chemotherapy response scores (CRS) after IDS.

**Results:** Among 65 enrolled patients, 39 NLHIPEC and 21 IV NACT patients underwent IDS. Grade 3-4 chemotherapy-related adverse effects were primarily hematological with no significant differences between two groups. The NLHIPEC group exhibited a higher proportion of CRS 3 (20.5% vs. 4.8%; P=0.000). R0 resection rates in IDS were 69.2% (NLHIPEC) and 66.7% (IV NACT). R2 resection occurred in 2.6% (NLHIPEC) and 14.3% (IV NACT) cases. No reoperations or postoperative deaths were reported, and complications were managed conservatively.

**Conclusions:** Combining NLHIPEC with IV NACT in treating ovarian cancer demonstrated safety and feasibility, with no increased chemotherapy-related adverse effects or postoperative complications. NLHIPEC improved tumor response to neoadjuvant chemotherapy, potentially enhancing progression-free survival (PFS). However, the final overall survival results are pending, determining if NLHIPEC combined with IV NACT is superior to IV NACT alone Keyword: high-grade serous ovarian/fallopian tubecarcinoma (HGSOC); paclitaxel; neoadjuvant chemotherapy;hyperthermic intraperitoneal chemotherapy (HIPEC); chemotherapy response score (CRS).

## Introduction

Ovarian cancer stands as the gynecologic malignancy with the highest mortality rate. Despite significant advances in targeted and immunotherapy in recent years, the overall 5-year survival rate remains below 50% [1]. High-grade serous cancer (HGSOC) predominates in this category [2]. HGSOC typically presents as asymptomatic and is challenging to diagnose at an early stage. Approximately 75% of HGSOC patients are diagnosed with advanced disease (FIGO IIIC-IVB stages), contributing to over 70% of all ovarian cancer-related deaths [2, 3].

Intraperitoneal dissemination serves as the primary mode of advanced HGSOC metastasis and is a key factor in treatment failure and recurrence [2]. There is evidence suggesting that combining intravenous and intraperitoneal chemotherapy can enhance and prolong patient survival [4, 5]. Nevertheless, the widespread adoption of this approach is impeded by catheter-related issues and the severe toxic side effects associated with intraperitoneal chemotherapy.

Intraperitoneal hyperthermic chemotherapy (HIPEC) represents an improved approach to intraperitoneal chemotherapy, and it has been employed in clinical practice for decades. The well-documented OVHIPEC-01 study validated the use of HIPEC in conjunction with interval debulking surgery (IDS) for enhancing the prognosis of patients with advanced epithelial ovarian cancer [6]. However, the effectiveness of HIPEC in the context of neoadjuvant chemotherapy remains uncertain. Recent retrospective studies have indicated that neoadjuvant HIPEC can enhance the chemotherapy response score (CRS) and reduce the recurrence rate among patients with advanced high-grade serous ovarian cancer [7]. Nevertheless, randomized trial data are currently lacking.

To address this gap in knowledge, we conducted a single-center, open-label Phase III randomized controlled Trial: The Impact of Paclitaxel-Based Neoadjuvant Laparoscopic Hyperthermic Intraperitoneal Chemotherapy (NLHIPEC) Followed by Sequential Intravenous Chemotherapy in Advanced High-Grade Serous Ovarian Cancer Patients –HIPEC for Ovarian Cancer in China (C-HOC Trial). Our aim was to investigate whether paclitaxel-based neoadjuvant HIPEC, when combined with intravenous neoadjuvant chemotherapy, offers an advantage over intravenous neoadjuvant chemotherapy alone in improving the NACT response score in patients with advanced HGSOC. Additionally, we explored whether the inclusion of HIPEC in neoadjuvant therapy led to increased adverse reactions and had a negative impact on IDS outcomes.

The primary endpoint of this trial was the difference in overall survival between the two groups. For this partial analysis, we compared the adverse effects of chemotherapy and postoperative complication rates after IDS between the two groups. Additionally, we assessed the immediate treatment efficacy by comparing the rate of pathological chemotherapy response score (CRS) after IDS.

## Methods

### Trial design

In a single-center, open-label, phase III trial, patients with FIGO stage IIIC-IVB HGSOC, who were evaluated with a FAGOTTI score ≥8 during laparoscopic exploration and were unable to undergo optimal cytoreduction (no visible disease (R0) or one or more residual tumors measuring 10 mm or less in diameter (R1) resection) in primary debulking surgery (PDS), were randomized into two groups in a 2:1 ratio. Randomization occurred at the time of laparoscopic exploration.

### Participants

#### Inclusion Criteria

This study encompasses newly diagnosed ovarian cancer patients falling within the age range of 18 to 75 years, exhibiting an Eastern Cooperative Oncology Group (ECOG) performance status score between 0 and 2. Included participants must not have undergone any prior anti-tumor therapies, including radiotherapy, chemotherapy, or targeted therapy. Eligible patients should have undergone a preoperative examination coupled with intraoperative exploration and evaluation, resulting in a diagnosis of International Federation of Gynecology and Obstetrics (FIGO) stage IIIC-IVB ovarian cancer. Additionally, participants must have attained a FAGOTTI score of at least 8 during laparoscopy, confirming the presence of High-Grade Serous Ovarian Cancer (HSGOC) through rapid pathology assessment. Moreover, individuals included in the study must exhibit sufficient bone marrow reserves and normal organ function, characterized by white blood cell counts of ≥3.5×10^9/L, neutrophil counts of ≥1.5×10^9/L, hemoglobin levels of ≥80g/L, and platelet counts of ≥80.0×10^9/L. Serum bilirubin, alanine aminotransferase (ALT), and aspartate aminotransferase (AST) levels should all be within the upper limits of normal. Likewise, urea nitrogen (BUN) and creatinine (Cr) levels should not exceed the upper limits of normal. Lastly, prospective participants must provide written informed consent to be included in the study.

#### Exclusion Criteria

This study excludes individuals with serious or uncontrolled medical and surgical conditions or acute infections. Pregnant or breastfeeding female patients are also ineligible for participation. Additionally, individuals with a history of gastrointestinal bleeding, perforation, intestinal obstruction, or related diseases are not included in this study.

#### Reasons for Dropout

Patients who were enrolled in the study were subject to dropout if they failed to adhere to the prescribed study protocol or voluntarily withdrew their consent for any personal reasons. Additionally, participants were removed from the study if they became unable to complete the planned treatment for any unforeseen circumstances or if they declined to undergo surgery at the same hospital as per the study requirements.

#### Data collection

The study was carried out and analyzed under the auspices of Department of Obstetrics and Gynecology at Ruijin Hospital. Oversight and monitoring of the study were conducted by the Clinical Research Center of Ruijin Hospital, the official body responsible for guiding and supervising various research endeavors within the hospital. Timely meetings were held to ensure adherence to protocol guidelines throughout the study’s implementation. This study received approval from the Ethics Committee of Ruijin Hospital and was registered on the Chinese Clinical Trial Registry platform (ChiCTR2000028894). Informed consent was obtained from all enrolled patients, beginning on September 2, 2019.

#### Interventions

##### Laparoscopic exploration

Patients underwent laparoscopic exploration under general anesthesia. Ascites, if present, were aspirated and measured. Suspected primary lesions or metastatic lesions were excised at a minimum of 2 points and sent for rapid pathology. FAGOTTI and Peritoneal Cancer Index scores (PCI) were calculated based on the exploration results.

##### HIPEC Treatment

After completing the initial endoscopic exploration, the experimental group underwent immediate HIPEC treatment under intraoperative general anesthesia. HIPEC was administered using the body cavity hyperthermic perfusion therapy system (Guangzhou Borui Medical Technology Co. LTDBR-TRG-II type) with paclitaxel 75 mg/m², normal saline 4000 ml, at a flow rate of 400-500 mL/min, and a temperature of 43±0.3°C for 60 minutes. The indwelling tubes included 2 inflow tubes and 2 outflow tubes. Temperature monitoring probes in both the inflow and outflow tubes ensured real-time intra-abdominal temperature monitoring, with a tolerance of ±0.3°C. Vital signs and tube patency were continuously monitored during thermal perfusion.

The experimental group (NLHIPEC group) received 1 cycle of neoadjuvant laparoscopic hyperthermic intraperitoneal chemotherapy (paclitaxel 75mg/m², 43±0.3°C, 60 minutes) followed by 3 cycles of paclitaxel + carboplatin, while the control group (IV NACT group) received only 3 cycles of intravenous neoadjuvant chemotherapy. In the NLHIPEC group an additional HIPEC was performed with the same chemotherapy regimen and dose, right after completing IDS.

##### Intravenous neoadjuvant chemotherapy (IV NACT)

Both the experimental and control groups initiated intravenous chemotherapy as soon as possible after exploratory surgery, administering IV NACT every 3 weeks for a total of 3 cycles. The experimental group (NLHIPEC group) received the following regimen: The first IV followed by HIPEC: paclitaxel 100 mg/m² on day 1 and carboplatin AUC=5 on day 2. The subsequent two regimens were: paclitaxel 175mg/m² on day 1 and carboplatin AUC=5 on day 2. The control group (IV NACT group) received only intravenous chemotherapy, with paclitaxel 175mg/m² on day 1 and carboplatin AUC=5 on day 2.

##### Interval Debulking Surgery (IDS)

After completing 3 cycles of chemotherapy, disease assessment was performed. IDS was performed in cases of CR/PR, while IDS was not considered if disease progression occurred during NACT. Open surgery under general anesthesia was performed, and the abdominal cavity was comprehensively investigated following a standardized pattern. FAGOTTI and PCI scores were determined based on the probe results. IDS was performed after exploration, with the aim of removing all visible lesions in the abdominal cavity. Surgical outcomes, blood loss, and perioperative blood transfusion were recorded, and all excised specimens were sent for pathological evaluation.

#### Outcomes

The initial primary endpoint of this phase III trial was to compare difference in overall survival between the two groups. Secondary endpoints included the IDS R0 resection rate, the Aletti score of IDS, surgical safety (length of stay after laparoscopic exploration and IDS, length of days from IV NACT after laparoscopic exploration, intraoperative blood loss, perioperative red blood cell transfusion), surgical complications, and NACT grade 3-4 adverse reactions (ARDs).

For this partial analysis, we compared the adverse effects of chemotherapy and postoperative complication rates after IDS between the two groups. Additionally, we assessed the immediate treatment efficacy by comparing the rate of pathological chemotherapy response score (CRS) after IDS.

CRS evaluates the pathological response to NACT in stage IIIC-IVB HGSOC patients, primarily based on omentum lesion retraction after neoadjuvant chemotherapy. CRS 1 indicates no or minimal tumor response, CRS 2 indicates a marked neoplastic response, and CRS 3 indicates complete or near-complete response. All patients underwent CRS scoring after IDS pathology confirmation by two pathologists.

#### Sample size

Assuming the control group has a 5-year survival rate of 38% and the experimental group has a 5-year survival rate of 60%, Significance level (α) = 0.05, Power (1 - β) = 0.8, Allocation ratio = 2:1, Estimated dropout rate = 10%, Estimated recruitment time = 2 years, It is calculated that the sample size for the experimental group is 90 cases, the control group is 45 cases, and the total sample size is 135 cases.

This is an Interim analysis of partial data for the reason describe above in the introduction. The data analysis cut-off time for this was set at the completion of surgery for the 60^th^ patient, who met the criteria for per-protocol (PP) analysis.

##### Recruitment

In the recruitment process, any ovarian cancer patient admitted to the center who met the predefined inclusion criteria was eligible for consideration and potential recruitment. This approach confirms that there was no selection bias during the recruitment phase, as all eligible patients were considered. The screening of patients was conducted by skilled clinicians at the designated center, and the principal investigators assumed responsibility for evaluating the pretreatment assessments and making enrollment decisions, ensuring a rigorous and unbiased recruitment process.

#### Randomisation

In our study, we employed a straightforward randomization approach without employing blocks or stratifying factors. The randomization process was carried out using a pre-established code. The generation of the random allocation sequence was overseen by a statistician at the Clinical Research Center, which serves as the central body responsible for supervising all clinical trials. The generation of the randomized code was accomplished using the Random Number Generators within the SPSS statistical software, with the initial seed value set to a reproducible fixed value. This process resulted in a randomized sequence designed for a 2:1 allocation, encompassing the total of 135 cases, with 90 cases experimental group, and 45 cases in the control group.

To maintain the integrity of the randomization, random numbers were placed inside sealed envelopes, each of which was sequentially numbered in accordance with the allocation sequence of the randomized numbers. These envelopes were subsequently opened in chronological order, corresponding to the admission sequence of the study subjects.

#### Blinding

A blinded statistician assumed responsibility for the randomized assignment of interventions, either to the experimental group (NLHIPEC group) or control group (IV NACT group). This assignment was executed through telephone contact or text messages, following the confirmation that the patient met the inclusion criteria and had provided informed consent. Importantly, both the patient and their caregivers were not blinded to the allocated intervention after assignment. However, outcome assessments were meticulously conducted by pathologists who were strictly blinded to the intervention group.

#### Statistical methods

For the analysis of the primary endpoint (CRS rate), we employed the intention-to-treat (ITT) population, comprising all patients who were randomly assigned to a treatment group. Postoperative morbidity and mortality, on the other hand, were analyzed within the per-protocol (PP) population, which consisted of patients who underwent surgery following the completion of all planned treatment.

Our statistical analysis was carried out using Statistical Package for Social Science (SPSS) version 22.0 for Windows, provided by SPSS, Inc. based in Chicago, Illinois. The normality of the data was assessed using the Kolmogorov-Smirnov Test. Continuous data were described using the median and range, while categorical data were presented as frequencies and percentages. To compare differences in rates between the two groups, the Fisher’s exact test was employed. All reported p-values are two-sided, and statistical significance was established at a threshold of less than 0.05. To ensure the robustness of our findings, all results underwent replication by two independent statisticians. Each statistician independently verified the results on two separate occasions, resulting in a total of four successful replications, thus enhancing the reliability of our statistical analyses.

## Results

This study commenced enrollment on September 2, 2019, and continued until March 3, 2023. A total of 65 patients enrolled, with 42 in the experimental group and 23 in the control group. Of these, 60 patients were included in the final analysis. Five patients were excluded (3 from the experimental group and 2 from the control group) for different reasons, including 3 patients not completing neoadjuvant chemotherapy as planned in our center, and 2 patients opting out of interval debulking surgery (Fig. 1). The final cohort comprised 39 cases in the experimental group and 21 cases in the control group. The phase III trial of this study is still going on; we present an interim analysis of partial data regarding the safety and immediate efficacy of both regimens. This analysis serves to determine the feasibility of proceeding with the planned phase III trial.

**Fig. 1.**
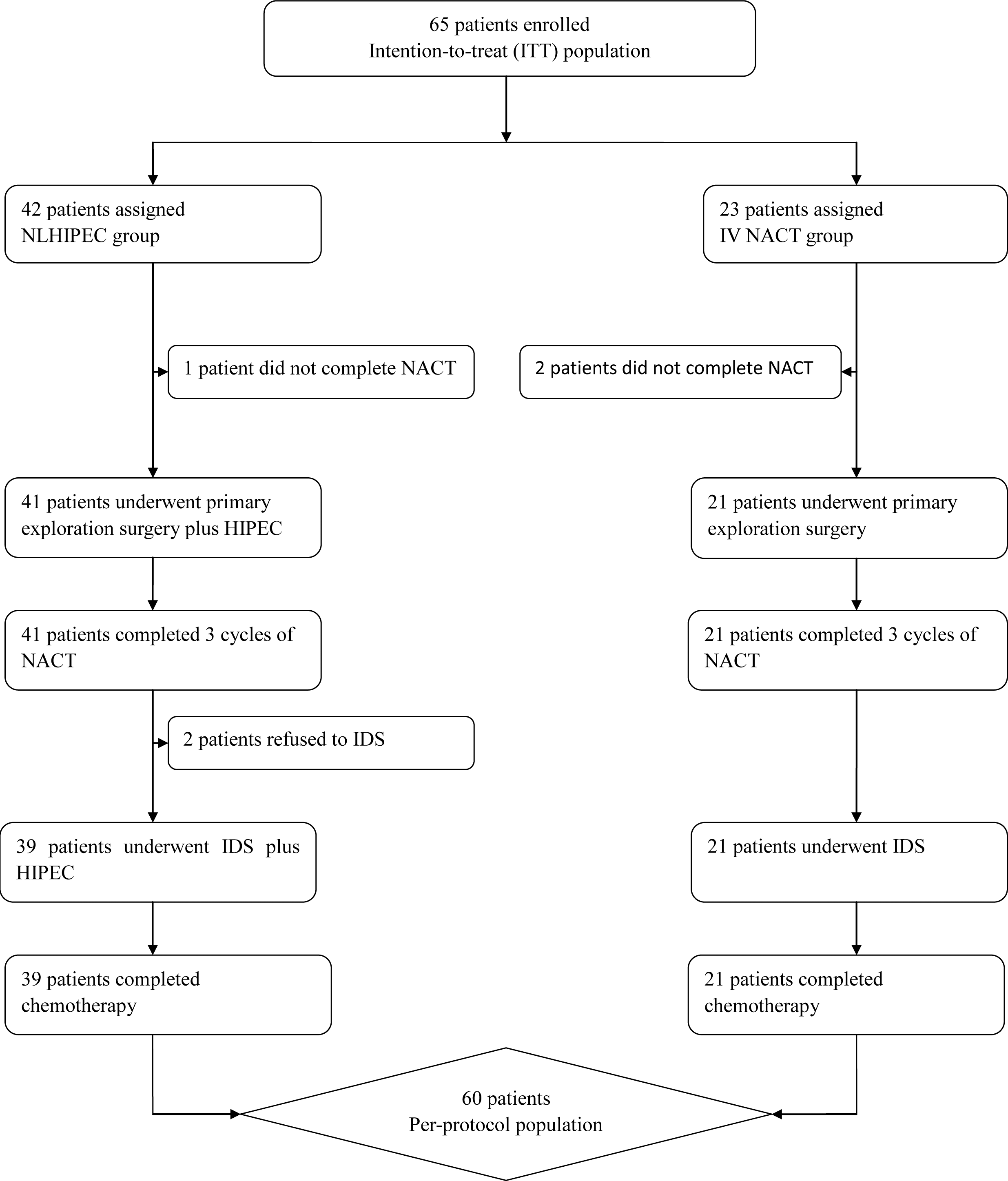
CONSORT diagram.

All patients were confirmed to have High-Grade Serous Ovarian Carcinoma (HGSOC) through postoperative paraffin pathology, which was consistent with intraoperative rapid pathology assessment. Moreover, all patients achieved either complete response (CR) or partial response (PR) after three cycles of NACT. The baseline characteristics of the enrolled patients are detailed in Table 1. Grade 3 to 4 adverse effects related to NACT chemotherapy primarily involved hematological issues, with no significant differences observed between the two groups (Table 2).

**Table 1.** Demographic.

| Variables |  | IV-NACT | NLHIPEC | P value |
| --- | --- | --- | --- | --- |
| Number of total cases |  | 21 | 39 | NA |
| Age (Years) | Median | 56 | 65 | 0.068 |
| BMI | <25 | 13(61.9) | 32(82.1) | 0.086 |
|  | >25 | 8(38.1) | 7(17.9) |  |
| ECOG | 0 | 17(81.0) | 31(79.5) | 0.892 |
|  | 1 | 4(19.0) | 8(20.5) |  |
| FIGO Stage | IIIC | 8(38.1) | 15(38.5) | 1.000* |
|  | IVA | 0 | 1(2.6) |  |
|  | IVB | 13(61.9) | 23(59.0) |  |
| Pretreatment CA 125 | Median | 1541.2 | 1785.5 | 0.932 |
| PCI score (exploration) | Median | 18 | 18 | 0.549 |
| BRCA mutation | None | 12(57.1) | 24(61.5) | 0.748* |
|  | BRCA1 | 4(19.0) | 9(23.1) |  |
|  | BRCA2 | 5(23.8) | 6(15.4) |  |

\*Fisher's exact test

**Table 2.**
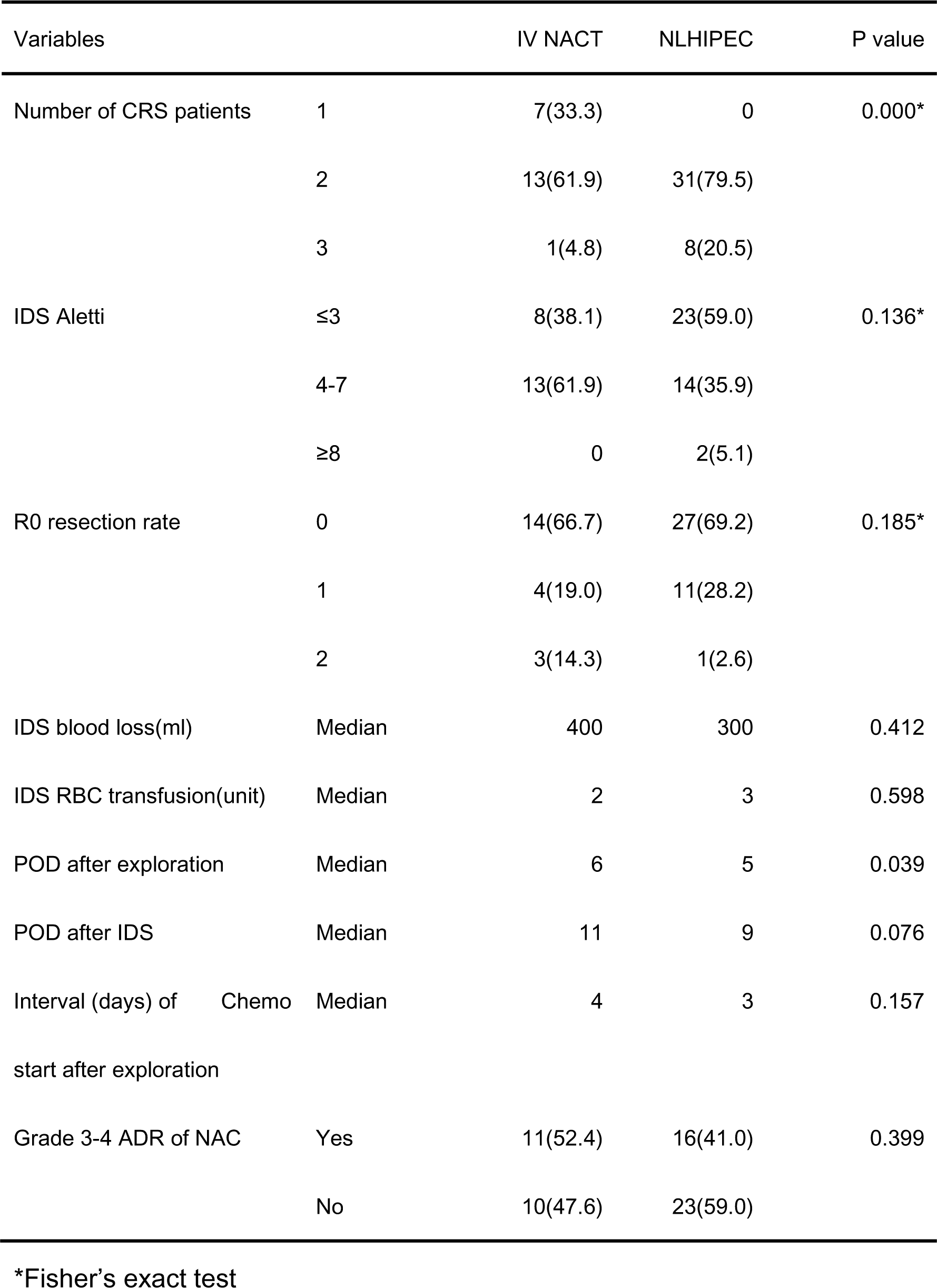
Treatment efficacy.

There were no reoperations for postoperative complications or fatalities in either group. No statistically significant differences were observed between the two groups in ALETTI scores, perioperative blood loss during IDS, the requirement for perioperative red blood cell transfusions, the time from the first intravenous chemotherapy after laparoscopic exploration, or the length of hospital stay following IDS. However, the experimental group did exhibit a shorter hospital stay following the initial laparoscopic exploration, as detailed in Table 2.

Complications associated with surgery included deep vein thrombosis of the lower limbs (DVT) in 1 case (experimental group), pulmonary embolism in 1 case (control group), and postoperative non-infectious fever in 1 case (experimental group). Postoperative intestinal obstruction occurred in 2 cases (1 in the experimental group and 1 in the control group), postoperative non-infectious fever occurred in 1 case (experimental group), and postoperative hemorrhage exceeding 1000ml occurred in 1 case (control group). All complications were successfully managed through conservative treatment.

Among the 39 patients in the experimental group, none underwent CRS1, while 31 cases (79.49%) underwent CRS2, and 6 cases (20.51%) achieved CRS3. In the control group consisting of 21 patients, 7 (33.33%) achieved CRS1, 14 (61.90%) achieved CRS2, and only 1 (4.76%) achieved CRS3. There was a statistically significant difference (p<0.05) in the rate of CRS between the two groups, as indicated in Table 2. No significant difference was observed in R0 resection rates between the two groups. R2 resection was performed in 1 case (2.6%) in the experimental group and 3 cases (14.3%) in the control group.

## Discussion

Currently, primary debulking surgery (PDS) combined with platinum-based chemotherapy stands as the primary treatment for High-Grade Serous Ovarian Carcinoma (HGSOC) [2, 11, 12]. However, in patients with advanced tumors, the extent of surgical intervention during PDS may be limited due to tumor spread, the need for extensive surgical procedures, and high surgical risks. This limitation often results in incomplete tumor cell reduction, leaving one or more residual lesions with a diameter exceeding 1 cm (classified as R2 residual disease). In such cases, interval debulking surgery (IDS) becomes an alternative option following neoadjuvant chemotherapy (NACT) with complete remission (CR) or partial remission (PR) achieved, as recommended by guidelines [11, 12]. Although the NACT+IDS approach remains a subject of debate, studies have demonstrated that when IDS achieves complete tumor cell reduction (defined as R0 resection, indicating no visible disease), the survival outcomes can be comparable to those of optimal cell reduction surgery (defined as R1 resection, where one or more residual tumors measure ≤1 cm in diameter) performed during PDS [13, 14]. Moreover, the IDS+NACT model is associated with lower surgical risks, reduced complication rates, and enhanced quality of life for advanced ovarian cancer patients [13, 14]. To further improve the efficacy of NACT and minimize surgical risks, novel approaches are needed.

Hyperthermic Intraperitoneal Chemotherapy (HIPEC) involves the continuous circulation of heated chemotherapy drugs within the abdominal cavity. Malignant tumors can experience irreversible damage at 43°C for 1 hour, while normal tissues can withstand temperatures up to 47°C for the same duration. Compared to intravenous chemotherapy, peritoneal perfusion offers advantages by bypassing the barrier effect of the peritoneum and directly and effectively targeting abdominal tumors [15, 16]. High temperatures can not only directly damage tumor cells but also induce tumor cell apoptosis through protein degeneration, angiogenesis inhibition, and changes in cell membrane permeability [17]. The activation of heat shock proteins, especially when combined with paclitaxel, can enhance drug toxicity and promote tumor cell apoptosis [18]. Moreover, HIPEC has been shown to induce apoptosis in distant metastatic lesions.

In our study, we employed the Chemotherapy Response Score (CRS) to assess the effectiveness of NACT. The CRS is recommended by the International Cancer Reporting Cooperation Organization and the European Society of Medical Oncology as a reliable prognostic tool for HGSOC patients [19, 20]. CRS3, in particular, has been associated with improved prognosis in HGSOC patients and can serve as an alternative indicator for Progression-Free Survival (PFS) [21]. While there was no difference in R0 resection rates between the two groups, the experimental group exhibited a significantly reduced R2 resection rate. It’s worth noting that the assessment of tumor reduction largely relies on subjective evaluation by surgeons and may entail some margin for error. Notably, there were no instances of reoperation or mortality in either group, and there was no significant difference in the proportion of patients undergoing complex surgery (as indicated by an ALETTI score >3) between the two groups. Therefore, paclitaxel-based HIPEC did not add complexity to IDS. Additionally, safety indicators such as length of hospital stay after IDS, time from intravenous NACT to laparoscopic exploration, intraoperative blood loss, and the incidence of perioperative red blood cell transfusion or grade 3-4 chemotherapy side effects did not significantly differ between the groups. Importantly, the experimental group even demonstrated a shorter hospital stay after the initial laparoscopic exploration, suggesting that the addition of neoadjuvant laparoscopic HIPEC (NLHIPEC) had no adverse impact on patient outcomes.

No differences in the status of Breast Cancer Susceptibility Genes (BRCA) mutations were observed between the two groups. It is widely recognized that patients with BRCA mutations exhibit greater responsiveness to platinum-based drugs [22] and can benefit from maintenance therapy with PARP inhibitors [23, 24], often resulting in improved prognosis. Our findings suggest that the addition of NLHIPEC may offer greater benefits to patients with wild-type BRCA.

Our paclitaxel-based NLHIPEC approach offers several advantages. Firstly, it utilizes a minimally invasive closed-mode HIPEC, which confines treatment to a controlled space, minimizing heat loss, preventing drug evaporation, and reducing the risk of drug contamination. Pharmacokinetic studies of cisplatin HIPEC have demonstrated that minimally invasive HIPEC, when compared to open surgery, enhances drug uptake in peritoneal tissue, with potential correlations to improved survival [25]. Secondly, we employed paclitaxel HIPEC instead of platinum-based drugs, leveraging paclitaxel’s favorable pharmacokinetics. Paclitaxel has a higher intraperitoneal-to-intravenous concentration ratio (AUC) compared to carboplatin (10:1) or cisplatin (20:1), leading to prolonged abdominal retention time. Furthermore, paclitaxel, in combination with hyperthermia, exhibits a synergistic effect, enhancing cytotoxicity and apoptosis of tumor cells [18]. The systemic toxicity of HIPEC is influenced by the pharmacokinetics and pharmacodynamics of the drugs used, as well as their dosage. Paclitaxel’s high molecular weight and hepatic metabolism result in minimal systemic toxicity, in contrast to cisplatin (molecular weight, MW 300.01 g/mol). Additionally, paclitaxel reduces the risk of kidney injury associated with cisplatin, similar to the observed benefits in OVHIPEC experiments. Clinical trials have indicated that paclitaxel HIPEC outperforms cisplatin and carboplatin in terms of Progression-Free Survival (PFS) and Overall Survival (OS) [27, 28]. Lastly, our study maintained a treatment temperature of 43°C, with strict temperature control provided by a body cavity hyperthermic perfusion therapy system. Research has shown that temperatures ranging from 41-43°C selectively destroy cancer cells without adversely affecting normal tissues [29, 30]. Studies on paclitaxel HIPEC have demonstrated that temperatures below 43°C enhance paclitaxel’s pro-apoptotic effects, with temperatures exceeding 43°C offering no significant additional benefit, confirming 43°C as the optimal and safe temperature. In our study, HIPEC was administered once under anesthesia supervision during surgery, reducing risks during perfusion therapy, as well as minimizing catheter-related complications and uneven drug distribution due to postoperative adhesions. Our study has certain limitations. First, as preliminary data analysis from a Phase III trial with a small sample size, we have not yet conducted long-term follow-ups for patients regarding Progression-Free Survival (PFS) and Overall Survival (OS). Additionally, due to economic constraints, our study included only the BRCA status of patients and did not account for Homologous Recombination Deficiency (HRD) status. Consequently, there may be a lack of efficacy evaluation for patients with wild-type BRCA and HRD mutations.

## Conclusion

Our study has provided evidence that the sequential administration of paclitaxel-based Neoadjuvant Laparoscopic Hyperthermic Intraperitoneal Chemotherapy (NLHIPEC) followed by intravenous neoadjuvant chemotherapy is associated with enhanced tumor response to neoadjuvant chemotherapy. Furthermore, this approach holds promise for improving Progression-Free Survival (PFS) while simplifying Interval Debulking Surgery (IDS). Importantly, the inclusion of NLHIPEC did not adversely affect the efficacy of NACT or the surgical procedures. Further research, including larger multi-center clinical trials and longer-term follow-ups for survival rate, is warranted to validate and expand upon these findings.

## Declaration

### Ethics Approval

This study received approval from the Ethics Committee of Ruijin Hospital and was registered on the Chinese Clinical Trial Registry platform (ChiCTR2000028894). The study adhered to the principles outlined in the Declaration of Helsinki (revised in 2013).

### Consent to Participate and Consent to Publish

Informed consent was diligently obtained from all enrolled patients.

### Author Contributions

WQ conceived and designed the study, collected patient data, and drafted the manuscript. LH and FWW contributed to the study’s design and critically reviewed manuscript drafts. All authors meet the criteria for publication, have read, and endorsed the final manuscript.

### Availability of Data and Materials

Datasets utilized and analyzed during this study are available from the corresponding author upon reasonable request.

## Acknowledgments

The authors extend their gratitude to all the healthcare professionals at the hospital who supported and facilitated the execution of this study.

